# Arterial Oxygen and Carbon Dioxide Tension and Acute Brain Injury in Extracorporeal Cardiopulmonary Resuscitation Patients: Analysis of the Extracorporeal Life Support Organization Registry

**DOI:** 10.1101/2022.03.10.22272203

**Authors:** Benjamin L Shou, Chin Siang Ong, Lavienraj Premraj, Patricia Brown, Joseph E. Tonna, Heidi J Dalton, Bo Soo Kim, Steven P Keller, Glenn JR Whitman, Sung-Min Cho

## Abstract

**Objective:** Acute brain injury remains common after extracorporeal cardiopulmonary resuscitation. Using a large international multicenter cohort, we investigated the impact of peri-cannulation arterial oxygen (PaO_2_) and carbon dioxide (PaCO_2_) on ABI occurrence.

**Design:** Retrospective cohort study.

**Setting:** Data in the Extracorporeal Life Support Organization Registry from 2009 to 2020.

**Patients:** Adult patients (≥18 years old) who underwent extracorporeal cardiopulmonary resuscitation.

**Interventions:** None.

**Measurements and Main Results:** Of 3,125 patients with extracorporeal cardiopulmonary resuscitation (median age=58, 69% male), 488 (16%) experienced at least one form of acute brain injury, which included ischemic stroke, intracranial hemorrhage, seizures, and brain death. 217 (7%) experienced ischemic stroke and 88 (3%) experienced intracranial hemorrhage. The registry collects two blood gas data pre- (6 hours before) and on- (24 hours after) extracorporeal membrane oxygenation (ECMO) cannulation. Blood gas parameters were classified as: hypoxia (<60mmHg), normoxia (60-119mmHg), and mild (120-199mmHg), moderate (200-299mmHg), and severe hyperoxia (≥300mmHg); hypocarbia (<35mmHg), normocarbia (35-44mmHg), mild (45-54mmHg) and severe hypercarbia (≥55mmHg). In multivariable logistic regression analysis, pre-ECMO hypoxia (aOR=1.46, 95%CI: 1.03-2.08, p=0.04) and on-ECMO severe hyperoxia (aOR=1.55, 95%CI: 1.02-2.36, p=0.04) were associated with composite ABI. Also, on-ECMO severe hyperoxia was associated with intracranial hemorrhage (aOR=1.88, 95%CI: 1.02-3.47, p=0.04) and in-hospital mortality (aOR=3.51, 95%CI: 1.98-6.22, p<0.001). Pre- and on-ECMO PaCO_2_ levels were not significantly associated with composite ABI or mortality, though mild hypercarbia pre- and on-ECMO were protective of ischemic stroke and intracranial hemorrhage, respectively.

**Conclusions:** Early severe hyperoxia (≥300mmHg) on ECMO was a significant risk factor for acute brain injury and mortality for patients undergoing extracorporeal cardiopulmonary resuscitation. Careful consideration should be given in early oxygen delivery in ECPR patients who are at risk of reperfusion injury.

## Introduction

Extracorporeal cardiopulmonary resuscitation (ECPR) combines venoarterial extracorporeal membrane oxygenation (VA-ECMO) and cardiopulmonary resuscitation (CPR) as a rescue therapy for patients with refractory cardiac arrest. Though ECPR use has rapidly increased(1) and may improve survival outcome compared to conventional CPR(2,3), acute brain injury (ABI) commonly occurs in as many as 27-32%(4,5) of patients, leading to poor neurological outcome(3,6). Primary ABI is caused by global brain ischemia during cardiac arrest. In contrast, secondary ABI occurs as a consequence of ECMO support; the immediate cerebral blood flow restoration results in reperfusion injury(7). Cerebral reperfusion injury may be compounded by aggressive oxygen therapy and acute changes in carbon dioxide during peri-cannulation period(7– 9). ABI is not only a significant cause of morbidity, but also portends an increase in mortality in ECMO patients(10,11).

Although the exact mechanisms behind ABI during ECMO support remain to be elucidated, acute changes in arterial carbon dioxide (PaCO_2_) and oxygen tension (PaO_2_) have plausible mechanisms to cause ABI. PaCO_2_ serves as a key regulator of cerebral autoregulation(12) and neuronal metabolic demand(13), and rapid decreases in PaCO_2_ may cause ABI by the combined effect of increased neuronal excitability with decreased cerebral blood flow. Though tissue hypoxia is widely known to be injurious, hyperoxia and resulting toxicity from increased free radical formation is also detrimental, especially after rapid reperfusion(14). Both PaCO_2_ and PaO_2_ have previously been associated with ABI and mortality in VA- and venovenous (VV-) ECMO patients, but these findings have not been well studied in ECPR patients(15). As clinical experience accumulates and ECPR becomes widely used, focused research on management of neurologic complications and on-ECMO care such as optimal oxygen and carbon dioxide level is imperative to improve neurological outcomes of ECPR patients; however, evidence regarding this is currently sparse.

In this study, we utilized the Extracorporeal Life Support Organization (ELSO) registry to examine the relationship between peri-cannulation PaCO_2_ and PaO_2_ levels on occurrence of ABI in patients receiving ECPR. We hypothesized that greater degree of peri-cannulation CO_2_ removal and severity of hyperoxia after ECMO initiation are associated with ABI.

## Materials and Methods

### Study Design and Population

This study was approved by the Johns Hopkins Institutional Review Board with a waiver of informed consent. The ELSO Registry is a voluntary database that collects clinical information and outcomes of ECMO support in adults and children from >500 member centers worldwide(16). The registry collects demographics, pre-ECMO medical diagnoses, hemodynamic and laboratory values before and during ECMO support, complications during ECMO support including ABI, and outcome such as survival to hospital discharge. Diagnosis and medical history are reported according to the International Classification of Diseases, 9th Edition and 10th Edition codes.

A retrospective analysis of ECPR patients in the ELSO database from January 2009 to December 2020 was performed. The inclusion criteria were 1) patients 18 years old and older; and 2) patients who received one run of ECMO support for ECPR indication. We excluded 1) patients who underwent multiple runs to avoid complexity and bias of confounding the outcome data; and 2) patients treated with non-ECPR VA-ECMO and VV-ECMO.

### Data Collection and Definitions

ECPR is defined in the ELSO registry as the rapid deployment of VA-ECMO in the setting of unsuccessful conventional cardiopulmonary resuscitation(17). The ELSO case report form collects hemodynamic and arterial blood gas (ABG) values before and after ECMO cannulation (“pre-ECMO” and “on-ECMO”, respectively). The pre-ECMO ABG value is the closest measurement to cannulation in the 6 hours before ECMO initiation. The on-ECMO value is the closest value to 24 hours after cannulation but could be taken at any time in hours 18-30 after cannulation. Delta PaO_2_ (rise in PaO_2_ over 24 hours) is defined as on-ECMO PaO_2_ minus pre-ECMO PaO_2_. Delta PaCO_2_ (change in PaCO_2_ over 24 hours) is defined as on-ECMO PaCO_2_ minus pre-ECMO PaCO_2_. Relative delta PaO_2_ and PaCO_2_ were defined as the delta value divided by the pre-ECMO blood gas value. Physiologically improbable data values under the following conditions were treated as erroneous values: pH>8.0 or <6.5, PaO_2_>760, and PaCO_2_>240 or <20 mmHg. PaO_2_ and PaCO_2_ categories were defined as following: hypoxia (<60 mmHg PaO_2_), normoxia (60-119) mmHg, mild hyperoxia (120-199) mmHg, moderate hyperoxia (200-299) mmHg, severe hyperoxia (>300 mmHg), hypocarbia (<35 mmHg PaCO_2_), normocarbia (35-44 mmHg), mild hypercarbia (45-54 mmHg), and severe hypercarbia (>55 mmHg). Numerical cutoffs for categories were determined based on standard clinical practice and prior literature(8).

Various ECMO complications were also included in the study. ECMO circuit mechanical failure includes any component or equipment failures which require intervention, such as replacement. Renal replacement therapy includes the use of any dialysis or hemofiltration while on ECMO. Hemolysis was defined as free plasma hemoglobin >50 mg/dL. Gastrointestinal (GI) hemorrhage included any upper or lower GI bleeding requiring packed red blood cell or whole blood transfusion, endoscopic intervention, or use of a hemostatic agent.

We defined composite acute brain injury (ABI) to include ischemic stroke, intracranial hemorrhage (ICH), diffuse brain ischemia, brain death, seizure, and ABI requiring neurosurgical intervention. In the ELSO database, ischemic stroke is defined as central nervous system (CNS) infarction as confirmed by ultrasound, computed tomography (CT), or magnetic resonance imaging (MRI). ICH is comprised of intra- or extra-parenchymal CNS hemorrhage as confirmed by ultrasound, CT, or MRI, or any CNS other hemorrhage, including intraventricular hemorrhage. Subtypes of ICH, such as intracerebral, subarachnoid, or subdural hematoma, were not available in the database. Seizure included those detected by clinical assessment or electroencephalogram. Those with ischemia, intraventricular hemorrhage, and neurosurgical intervention, which were added as variables to the ELSO database in 2019 and after, were included as part of the ABI group.

### Outcomes

The primary outcome was composite ABI. The secondary outcomes were ischemic stroke, ICH, and mortality.

### Statistical Analysis

Continuous variables were expressed as medians with interquartile range (IQR). Categorical variables were expressed as frequencies with percentages. Wilcoxon rank-sum and Pearson’s chi-squared tests were used to compare continuous and categorical variables, respectively. Cuzick’s non-parametric test was used to assess for trends over time(18). P values < 0.05 were considered statistically significant.

Univariable and multivariable logistic regression were performed to determine associations between PaO_2_/PaCO_2_ metrics and 1) ABI, 2) ischemic stroke, 3) ICH, and 4) mortality. Variables which had a p<0.05 by univariable analysis were included in the initial multivariable regression model. Different permutations of PaO_2_/PaCO_2_ metrics were added to assess improvements in model performance. PaO_2_/PaCO_2_ metrics included in different models were categorized PaO_2_/PaCO_2_ values, as above, PaO_2_/PaCO_2_ as a continuous variable, delta, and relative delta PaO_2_/PaCO_2_. The models with the lowest Akaike’s information criterion (AIC) were selected as the final models. The final model for ABI (primary outcome) was adjusted for the following pre-ECMO characteristics based on these variables being statistically significant in univariable regression: age, race, pre-ECMO pH and lactate, requirement of renal replacement therapy, hemorrhagic hemolysis, and on-ECMO arrythmia. Adjusted odds ratios (aOR) were presented with 95% confidence intervals (CIs). Statistical significance was set at p<0.05. All analyses were carried out in STATA 17 (StataCorp, LLP, College Station, TX).

## Results

### Incidence and Mortality of ABIs

Of 3,125 patients (median age=58, 69% male) who underwent ECPR intervention, 488 (16%) experienced ABI. The most common type of ABI was ischemic stroke (n=217, 7%), brain death (n=183, 6%), ICH (n=88, 3%), and seizures (n=69, 2%). Except for a decrease in 2020, the use of ECPR has steadily increased over time from 36 patients in 2009 to 677 in 2019 (p-trend=0.003) (**Figure 1A**). The incidence of composite ABI among ECPR patients has decreased over time (p-trend=0.02). However, the incidence of ischemic stroke has increased while ICH has remained stable (**Figure 1B**). Those with ABI (88% vs. 65%), ischemic stroke (84% vs. 68%), and ICH (89% vs. 68%) had higher mortality compared to those without (p<0.001 for all).

**Figure 1.**
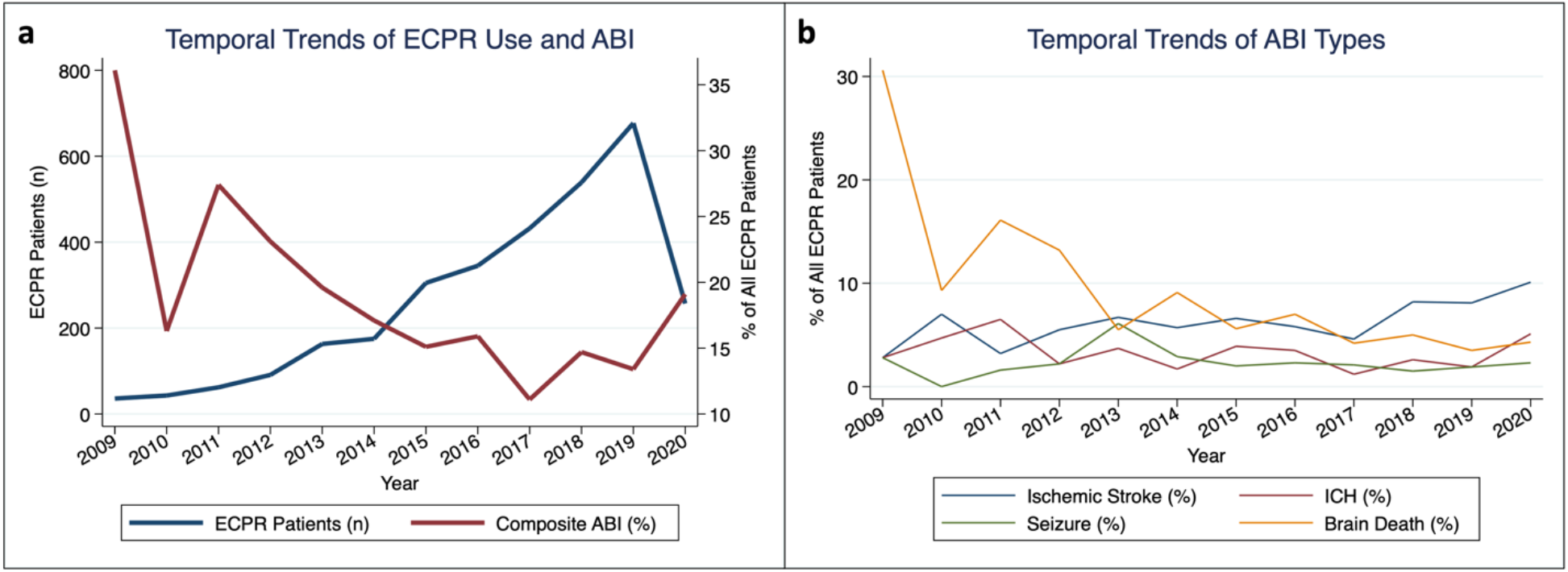
Trends in the (a) utilization of extracorporeal cardiopulmonary resuscitation (ECPR) and incidence of acute brain injury (ABI), and (b) for the incidence of individual ABI types. Trends were assessed using Cuzick’s non-parametric trend test. Annual ECPR volume has increased (p-trend=0.003) while incidence of ABI has decreased (p-trend=0.023). Incidence of brain death decreased and ischemic stroke increased, while incidence of ICH and seizure have not changed over time.

### Arterial Oxygen Tension and ABI

Patients with ABI had a lower median pre-ECMO PaO_2_ compared to those without (73 mmHg vs. 77, p=0.02) (**Table 1)**. Following ECMO initiation, those with ABI had a higher PaO_2_ (156 vs. 133 mmHg, p<0.001) and delta PaO_2_ (+84 vs. +50 mmHg, p<0.001), and were more likely to have moderate (13% vs. 10%, p=0.003) or severe hyperoxia (22% vs. 15%, p=0.003). Additionally, ABI patients had a higher relative delta PaO_2_ (+112% vs. +67%, p<0.001). Distributions of on-ECMO PaO_2_ and PaCO_2_ are plotted in **Figure 2**. There was a U-shaped relationship between oxygenation status and frequency of composite ABI, ischemic stroke, and ICH (**Figures S1A, S1C, S1E**). Compared to other types of ABI, those with ICH experienced the largest increase in PaO_2_ as well as highest on-ECMO PaO_2_ (**Figure S2**).

**Table 1.** Demographics, clinical characteristics, and extracorporeal membrane oxygenation characteristics for extracorporeal cardiopulmonary resuscitation patients who had acute brain injury (ABI) versus those who did not.

|  | Total<br>(n=3,125) | No ABI<br>(n=2,637, 84.4%) | ABI<br>(n=488, 15.6%) | p value |
| --- | --- | --- | --- | --- |
| Age (years) | 58 (47-67) | 59 (48-67) | 54 (43-64) | <0.001 |
| Female sex | 961 (31%) | 814 (31%) | 147 (30%) | 0.79 |
| Weight (kg) | 81 (70-97) | 80 (70-96) | 84 (70-100) | 0.10 |
| Race |  |  |  | 0.03 |
| White | 1,789 (57%) | 1,541 (58%) | 248 (51%) |  |
| Asian | 627 (20%) | 519 (20%) | 108 (22%) |  |
| Black | 274 (9%) | 221 (8%) | 53 (11%) |  |
| Hispanic | 114 (4%) | 92 (3%) | 22 (5%) |  |
| Other | 235 (8%) | 191 (7%) | 44 (9%) |  |
| Comorbidity |  |  |  |  |
| Diabetes | 304 (10%) | 253 (10%) | 51 (10%) | 0.56 |
| Hypertension | 234 (7%) | 189 (7%) | 45 (9%) | 0.11 |
| Atrial fibrillation | 132 (4%) | 107 (4%) | 25 (5%) | 0.28 |
| Cardiomyopathy | 280 (9%) | 235 (9%) | 45 (9%) | 0.83 |
| COPD | 96 (3%) | 79 (3%) | 17 (3%) | 0.57 |
| Pre-ECMO mean blood pressure (mmHg) | 57 (41-72) | 57 (42-71) | 56 (40-75) | 0.87 |
| **Pre-ECMO ABG** |  |  |  |  |
| pH | 7.17 (7.02-7.29) | 7.18 (7.02-7.30) | 7.11 (6.97-7.25) | <0.001 |
| HCO<sub>3</sub><sup>-</sup> (mEq/L) | 18 (13-22) | 18 (14-22) | 17 (13-21) | 0.003 |
| PaO<sub>2</sub> | 76 (57-112) | 77 (57-112) | 73 (54-108) | 0.018 |
| PaCO<sub>2</sub> | 49 (38-63) | 48 (38-62) | 51 (40-66) | 0.013 |
| Lactate (mmol/L) | 10.0 (5.6-14.3) | 9.9 (5.3-14.0) | 11.6 (7.4-15.4) | <0.001 |
| **Pre-ECMO ABG O<sub>2</sub> Categories** |  |  |  | 0.0 |
| Hypoxia (<60 mmHg) | 900 (29%) | 737 (28%) | 163 (33%) |  |
| Normoxia (60-119 mmHg) | 1,526 (49%) | 1,300 (49%) | 226 (46%) |  |
| Mild hyperoxia (120-199 mmHg) | 451 (14%) | 382 (14%) | 69 (14%) |  |
| Moderate hyperoxia (200-299 mmHg) | 248 (8%) | 218 (8%) | 30 (6%) |  |
| Severe hyperoxia (≥300 mmHg) | 0 (0%) | 0 (0%) | 0 (0%) |  |
| **Pre-ECMO ABG CO<sub>2</sub> Categories** |  |  |  | <0.001 |
| Hypocarbica (<35 mmHg) | 506 (16%) | 431 (16%) | 75 (15%) |  |
| Normocarbica (35-44 mmHg) | 760 (24%) | 645 (24%) | 115 (24%) |  |
| Mild hypercarbica (45-54 mmHg) | 665 (21%) | 592 (22%) | 73 (15%) |  |
| Severe hypercarbica (≥55 mmHg) | 1,194 (38%) | 969 (37%) | 225 (46%) |  |
| **Post-ECMO ABG (24 hrs)** |  |  |  |  |
| pH | 7.41 (7.35-7.46) | 7.41 (7.35-7.46) | 7.40 (7.35-7.45) | 0.64 |
| HCO<sub>3</sub><sup>-</sup> (mEq/L) | 24 (20-26) | 24 (20-26) | 23 (20-26) | 0.70 |
| PaO<sub>2</sub> | 138 (89-257) | 133 (88-250) | 156 (99-305) | <0.001 |
| PaCO<sub>2</sub> | 37 (33-42) | 37 (33-42) | 37 (32-42) | 0.97 |
| Lactate (mmol/L) | 3.4 (1.8-7.2) | 3.3 (1.8-7.2) | 3.8 (2.1-7.0) | 0.21 |
| **Post-ECMO ABG O<sub>2</sub> Categories** |  |  |  | 0.003 |
| Hypoxia (<60 mmHg) | 130 (4%) | 104 (4%) | 26 (5%) |  |
| Normoxia (60-119 mmHg) | 904 (29%) | 779 (30%) | 125 (26%) |  |
| Mild hyperoxia (120-199 mmHg) | 583 (19%) | 494 (19%) | 89 (18%) |  |
| Moderate hyperoxia (200-299 mmHg) | 336 (11%) | 274 (10%) | 62 (13%) |  |
| Severe hyperoxia (≥300 mmHg) | 501 (16%) | 394 (15%) | 107 (22%) |  |
| **Post-ECMO ABG CO<sub>2</sub> Categories** |  |  |  | 0.97 |
| Hypocarbica (<35 mmHg) | 831 (27%) | 692 (26%) | 139 (28%) |  |
| Normocarbica (35-44 mmHg) | 1,228 (39%) | 1,020 (39%) | 208 (43%) |  |
| Mild hypercarbia (45-54 mmHg) | 321 (10%) | 270 (10%) | 51 (10%) |  |
| Severe hypercarbia (≥55 mmHg) | 58 (2%) | 49 (2%) | 9 (2%) |  |
| Delta PaO<sub>2</sub> (mmHg) | 53 (1-171) | 50 (0-157) | 84 (14-216) | <0.001 |
| Relative delta PaO<sub>2</sub> (%) | 73 (1-231) | 67 (0-210) | 112 (18-314) | <0.001 |
| Delta PaCO<sub>2</sub> (mmHg) | -11 (-25 - -1) | -11 (-24 - -1) | -12 (-28 - -2) | 0.04 |
| Relative delta PaCO<sub>2</sub> (%) | -23 (-41 - -2) | -23 (-41 - -2) | -26 (-43 - -5) | 0.080 |
| Days on ECMO support | 3.0 (1.0-5.9) | 2.9 (0.9-5.9) | 3.2 (1.5-6.0) | <0.001 |
| Additional mechanical support device | 512 (16%) | 429 (16%) | 83 (17%) | 0.69 |
| ECMO complications |  |  |  |  |
| ECMO circuit mechanical failure | 435 (14%) | 356 (14%) | 79 (16%) | 0.12 |
| Renal replacement therapy | 834 (27%) | 648 (25%) | 186 (38%) | <0.001 |
| Glucose < 40 mg/L | 21 (1%) | 13 (0%) | 8 (2%) | 0.004 |
| Hemolysis | 82 (3%) | 62 (2%) | 20 (4%) | 0.03 |
| Cardiac arrhythmia | 455 (15%) | 364 (14%) | 91 (19%) | 0.005 |
| Gastrointestinal hemorrhage | 151 (5%) | 124 (5%) | 27 (6%) | 0.43 |
| Disseminated intravascular coagulation | 62 (2%) | 48 (2%) | 14 (3%) | 0.13 |
| **Outcomes** |  |  |  |  |
| Survival | 980 (31%) | 922 (35%) | 58 (12%) | <0.001 |

Continuous values are represented as # (interquartile range) and categorical variables are represented as n (%). Blood gasses have units of mmHg unless otherwise noted. Abbreviations – COPD: chronic obstructive pulmonary disease; ECPR: extracorporeal cardiopulmonary resuscitation; ECMO: extracorporeal membrane oxygenation; ABG: arterial blood gas; PaO<sub>2</sub>: arterial oxygen level; PaCO<sub>2</sub>: arterial carbon dioxide level. “Additional mechanical support device” includes left and right ventricular assist devices (VAD), intra-aortic balloon pumps, and percutaneous VADs. Delta PaO<sub>2</sub> = Post-ECLS PaO<sub>2</sub> minus Pre-ECLS PaO<sub>2</sub>. Delta PaCO<sub>2</sub> = Post-ECLS PaCO<sub>2</sub> minus Pre-ECLS PaCO<sub>2</sub>. “Other” race is defined as a separate category in the ELSO database.
Fields with physiologically improbable values were treated as missing: pH>8 or <6.5, PaO<sub>2</sub>>760, and PaCO<sub>2</sub>>240 or <20 mmHg. Missing variables for some variables result in different denominators, listed here: male sex (n=3,104), weight (n=3,032), race (n=3,039), pre-ECLS pH (n=3,113), HCO<sub>3</sub><sup>-</sup> (n=2,965), and lactate (n=1,592), post-ECLS pH (n=2,458), HCO<sub>3</sub><sup>-</sup> (n=2,421), PaO<sub>2</sub> (n=2,454), PaCO<sub>2</sub> (n=2,438), and lactate (n=1,405), delta PaO<sub>2</sub> (n=2,454), delta PaCO<sub>2</sub> (n=2,438), and pre-ECLS mean blood pressure (n=1,971).

**Figure 2.**
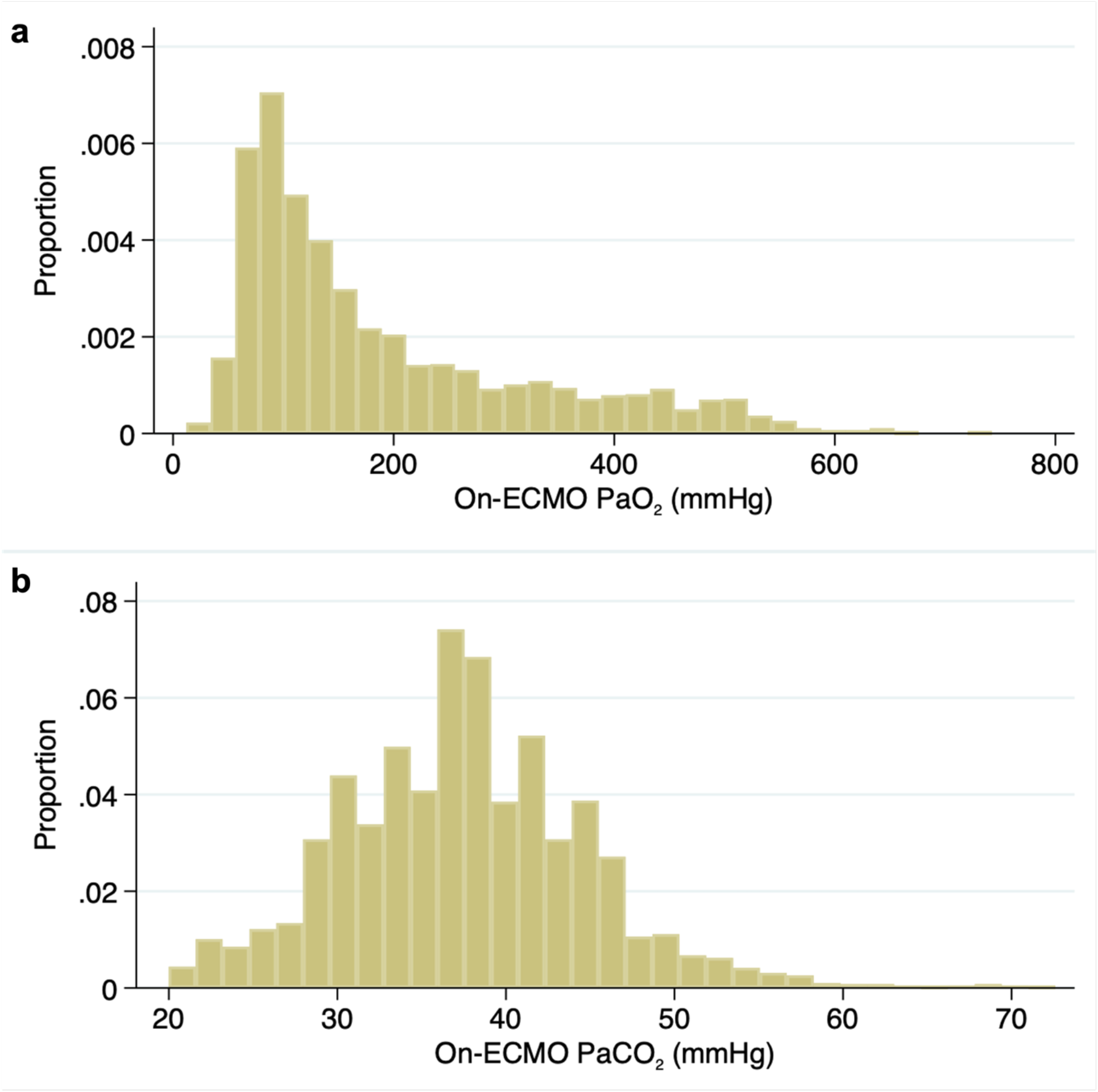
Histograms for (a) on-ECMO PaO_2_ and (b) on-ECMO PaCO_2_. Abbreviations – ECMO: extracorporeal membrane oxygenation.

### Arterial Carbon Dioxide Tension and ABI

Those with ABI had a higher pre-ECMO PaCO_2_ (51 vs. 48 mmHg, p=0.01), were more likely to have severe hypercarbia (46% vs. 37%, p<0.001), and less likely to have mild hypercarbia (15% vs. 22%, p<0.001) prior to ECMO initiation (**Table 1**). There were no differences in PaCO_2_ between those with and without ABI after ECMO initiation, however, the ABI group had a marginally larger delta PaCO_2_ (−12 vs. -11 mmHg, p=0.04).

### Risk Factors of ABI

#### Composite ABI

Differences in baseline characteristics, pre-ECMO, and ECMO variables between those with and without composite ABI are shown in **Table 1**. In the final multivariable model for composite ABI, lower pre-ECMO pH (aOR=1.13 per 0.1 unit drop, 95% CI: 1.03-1.25, p=0.01), pre-ECMO hypoxia (aOR=1.46, 95% CI: 1.03-2.08, p=0.04), on-ECMO severe hyperoxia (aOR=1.55, 95% CI: 1.02-2.36, p=0.04), and renal replacement therapy (aOR=1.70, 1.22-2.37, p=0.002) were significant risk factors of developing composite ABI (**Table 2, Figure 3A**). Delta PaO_2_ and delta PaCO_2_, and relative deltas, were not significantly associated with ABI.

**Table 2.** Factors associated with composite acute brain injury by multivariable logistic regression. C-statistic for model is 0.67. Variables with a p<0.05 are bolded.

|  | Multivariable |  |  |
| --- | --- | --- | --- |
|  | aOR | 95% CI | p value |
| Age (by 10 years) | 0.92 | 0.82-1.02 | 0.11 |
| Race |  |  |  |
| White | Reference |  |  |
| Black | 1.55 | 0.92-2.62 | 0.10 |
| Hispanic | 1.79 | 0.83-3.90 | 0.14 |
| Asian | 0.91 | 0.58-1.41 | 0.66 |
| Other | 1.54 | 0.98-2.42 | 0.06 |
| **Decreasing pH (per 0.1 units), pre-ECMO** | **1.13** | **1.03-1.25** | **0.01** |
| Lactate, pre-ECMO | 1.01 | 0.99-1.04 | 0.34 |
| PaO<sub>2</sub>, pre-ECMO |  |  |  |
| **Hypoxia (<60 mmHg)** | **1.46** | **1.03-2.08** | **0.04** |
| Normoxia (60-120 mmHg) | Reference |  |  |
| Mild hyperoxia (120-199 mmHg) | 1.13 | 0.71-1.80 | 0.62 |
| Moderate hyperoxia (200-299 mmHg) | 0.86 | 0.45-1.63 | 0.64 |
| PaCO<sub>2</sub>, pre-ECMO |  |  |  |
| Hypocarbica (<35 mmHg) | 0.86 | 0.50-1.48 | 0.59 |
| Normocarbica (35-44 mmHg) | Reference |  |  |
| Mild hypercarbica (45-54 mmHg) | 0.72 | 0.44-1.17 | 0.18 |
| Severe hypercarbica (≥55 mmHg) | 0.94 | 0.61-1.45 | 0.76 |
| PaO<sub>2</sub>, on-ECMO |  |  |  |
| Hypoxia (<60 mmHg) | 1.32 | 0.66-2.66 | 0.43 |
| Normoxia (60-120 mmHg) | Reference |  |  |
| Mild hyperoxia (120-199 mmHg) | 1.28 | 0.85-1.91 | 0.24 |
| Moderate hyperoxia (200-299 mmHg) | 1.34 | 0.83-2.17 | 0.24 |
| **Severe hyperoxia (≥300 mmHg)** | **1.55** | **1.02-2.36** | **0.04** |
| **Renal replacement therapy** | **1.70** | **1.22-2.37** | **0.002** |
| Hemolysis | 1.47 | 0.64-3.35 | 0.36 |
| Arrhythmia | 1.34 | 0.89-2.04 | 0.17 |

Abbreviations – ECMO: extracorporeal membrane oxygenation. PaO<sub>2</sub>: arterial oxygen level; PaCO<sub>2</sub>: arterial carbon dioxide level. Addition of delta PaO<sub>2</sub> and CO<sub>2</sub> parameters (both raw values and relative percentages) degraded model performance compared to using only the categorized variables and were consequently excluded from the final model.

**Figure 3.**
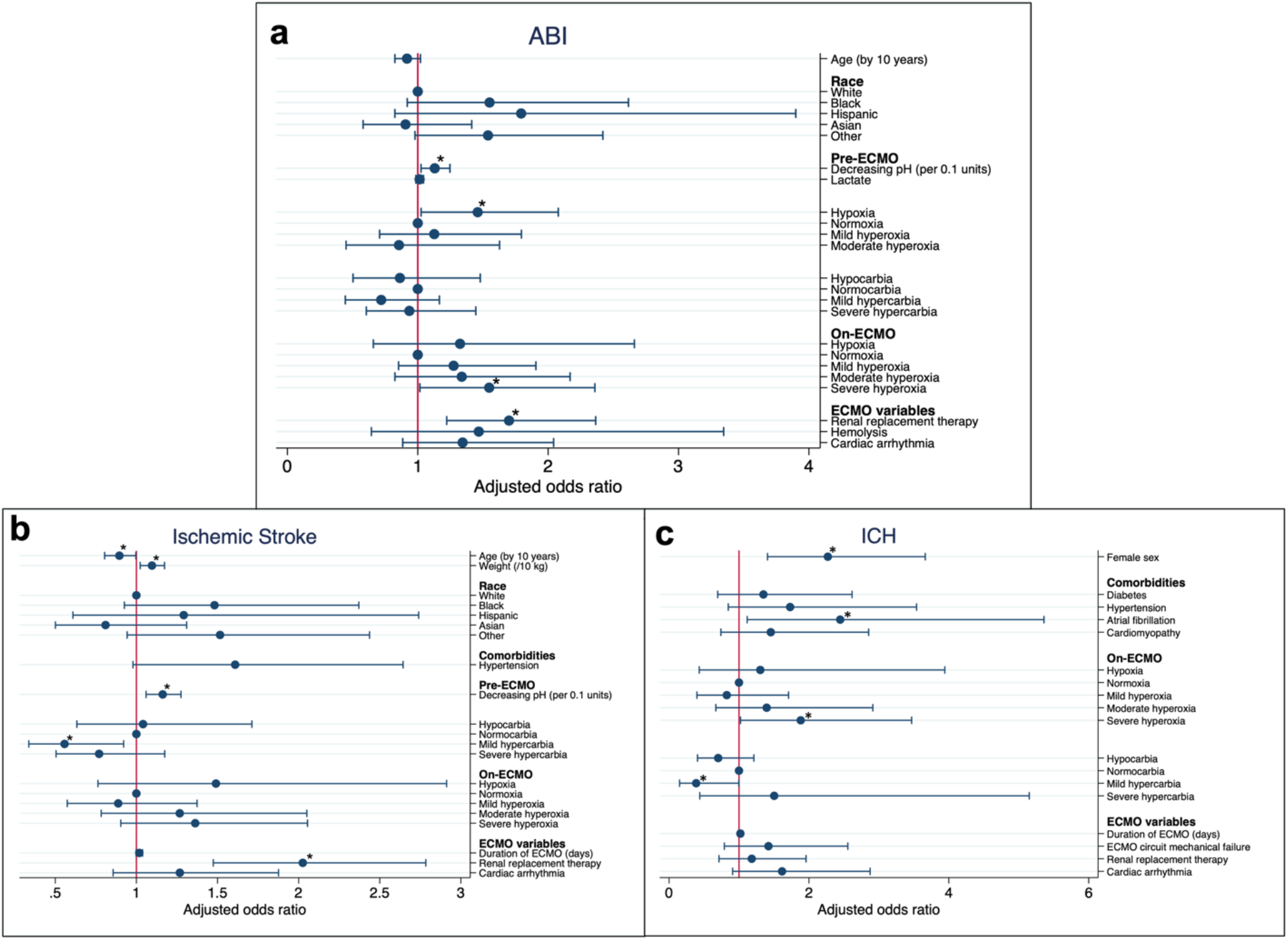
Forest plot for multivariable model of (a) acute brain injury (ABI), (c) ischemic stroke, and (c) intracranial hemorrhage (ICH). Dot represents adjusted odds ratio and brackets represent the 95% confidence intervals. Asterisks mark variables with a p<0.05.

#### Ischemic stroke

We separately analyzed secondary outcomes, ischemic stroke and ICH. In a multivariable logistic regression model for ischemic stroke (**Figure 3B, Table S1**), older age (aOR=0.90 for every 10 year age increase, 95% CI: 0.80-0.99, p=0.04) and pre-ECMO mild hypercarbia (aOR=0.56, 95% CI: 0.34-0.92, p=0.02) were protective against ischemic stroke. In the same model, higher weight (for every 10 kg increase, aOR=1.10, 95% CI: 1.02-1.17, p=0.008), lower pre-ECMO pH (aOR=1.16 per 0.1 unit drop, 95% CI: 1.06-1.28, p=0.001), and renal replacement therapy during ECMO (aOR=2.03, 95% CI: 1.47-2.78, p<0.001) were associated with ischemic stroke.

#### ICH

By multivariable logistic regression, (**Figure 3C, Table S2**), female sex (aOR=2.27, 95% CI: 1.41-3.66, p=0.001), atrial fibrillation (aOR=2.45, 95% CI: 1.12-5.36, p=0.03), and on-ECMO severe hyperoxia (aOR=1.88, 95% CI: 1.02-3.47, p=0.04) were associated with ICH. On-ECMO mild hypercarbia (aOR=0.39, 95% CI: 0.15-1.00, p=0.05) was protective against ICH.

### Risk Factors of Mortality

There was a U-shaped relationship between both on-ECMO PaO_2_ and delta PaCO_2_ with mortality (**Figure S3**). In a multivariable model, those who had ABI had more than a four-fold increased adjusted odds of mortality (aOR=4.18, 95% CI: 2.28-7.64, p<0.001) (**Figure S4, Table S3**). Increasing age (aOR=1.28 for every 10 year increase, 95% CI: 1.13-1.45, p<0.001) and weight (for every 10 kg increase, aOR=1.08, 95%CI: 1.00-1.17, p=0.05), as well as on-ECMO lactate (aOR=1.12, 95% CI: 1.07-1.18, p<0.001), lower pre-ECMO mean blood pressure (for every 10 mmHg decrease, aOR=1.15, 95% CI: 1.07-1.24, p<0.001) and lower on-ECMO pH (aOR=1.29 per 0.1 unit drop, 95% CI: 1.05-1.59, p=0.02) were associated with mortality. Severe hyperoxia on-ECMO was strongly associated with mortality (aOR=3.51, 95% CI: 1.98-6.22, p<0.001), as were ECMO circuit mechanical failure (aOR=2.17, 95% CI: 1.17-4.01, p=0.01), renal replacement therapy (aOR=2.24, 95% CI: 1.44-3.48, p<0.001), and on-ECMO arrythmia (aOR=2.56, 95% CI: 1.47-4.45, p=0.001). Similar to the ABI models, delta PaO_2_ and delta PaCO_2_, and relative deltas, were not significantly associated with mortality.

## Discussion

Our analysis of 3,125 ECPR patients within the ELSO registry revealed that 16% experienced at least one type of ABI. Ischemic stroke (7%) was the most common type of ABI, followed by brain death (6%), ICH (3%) and seizure (2%). ABI is a consequential and common occurrence in patients receiving ECPR: a meta-analysis of 78 studies (2008-2019) showed similar prevalence of ischemic stroke, brain death and ICH, though overall prevalence of ABI was higher in that study (27%)(4). We observed an 18-fold increase in ECPR use between 2009 and 2019, as increasing evidence accumulates for the benefit of ECPR over conventional CPR(7). Rates of composite ABI and brain death decreased while frequency of cerebrovascular events, such as ischemic stroke and intracranial hemorrhage, remained unchanged (**Figure 1**). Since ECPR survival has improved in the modern era(4), these epidemiological data may point to better selection of patients who may benefit from ECPR as well as more standardized practice and experience(19) while the prevention and treatment of on-ECMO cerebrovascular complications remains a challenge.

In cardiac arrest patients, early and prolonged hyperoxia (PaO_2_>300 mmHg) is associated with mortality and poor neurological outcome after both conventional CPR(20) and during VA-ECMO (non-ECPR specific)(8). A retrospective analysis of adult ECPR patients within the ELSO database from 2010 to 2015 showed that moderate hyperoxia (PaO_2_ 101-300 mmHg), but not severe hyperoxia (PaO_2_>300 mmHg), was associated with increased mortality(21). The current study is the first to utilize a large international ECPR cohort to demonstrate early severe hyperoxia is a strong risk factor of ABI, which contributes to significant mortality and morbidity after ECMO. Severe hyperoxia likely exacerbates reperfusion injury in ECPR by providing greater substrate for toxic free radical formation(14) and subsequent derangements like neuronal metabolic failure(22) and a pro-inflammatory state(23). Given the clinical evidence and mechanistic plausibility of ABI caused by hyperoxia, we need clinical trials to determine optimal oxygenation targets within the first 24 hours after ECPR where ischemic reperfusion injury risk is highest.

Pre-ECMO hypoxia was also associated with increased risk of composite ABI but not with ischemic stroke or ICH individually. Cardiac arrest results in temporary global anoxia and low cerebral blood flow, so pre-ECMO hypoxia likely mediates hypoxic ischemic brain injury (HIBI) and subsequent brain death rather than increasing the risk of cerebrovascular events, which are likely on-ECMO complications. A meta-analysis of ECPR studies reported HIBI as the most common type of ABI (23%) by a substantial margin(4). Though HIBI is not explicitly recorded in the ELSO registry, our cohort had similarly high rates of brain death (38% of all ABI), majority of which may be caused by HIBI.

The role of hypercarbia in ECMO is contentious. On one hand, an analysis of VA-ECMO patients in the ELSO registry, PaCO_2_ was associated with increased mortality at tensions <30 mmHg and >60 mmHg(24). On the other hand, Munshi et al. reported that mild hypercarbia (PaCO_2_>45 mmHg) was inversely associated with mortality (OR=0.78, 95% CI=0.32-1.93) in ECPR patients(21). In this study, did not find any associations between PaCO_2_ and mortality. Regarding ABI, on-ECMO mild hypercarbia (45-54 mmHg) was protective against ICH while pre-ECMO mild hypercarbia protected against ischemic stroke. PaCO_2_ may play a distinct physiological role in ECPR versus VA-ECMO. Hypercarbia is known to increase cerebral blood flow and, in a phase II randomized controlled trial, was associated with lower serum levels of neuronal and glial injury biomarkers (serum neuron specific enolase and S100b) 24 hours post-cardiac arrest compared to normoxia in post-cardiac arrest patients(25). Thus, mild hypercarbia in peri-cannulation period may attenuate the risk of ABI.

Despite delta PaCO_2_ in ABI patients being marginally higher than in non-ABI patients in univariate analysis, delta PaCO_2_ was not significantly associated with ABI or mortality after adjustment. Neither sensitivity analysis nor categorization using ordinal analyses of PaCO_2_ yielded meaningful trends. Prior studies with VA-ECMO patients are equivocal. With the benefit of protocolized neuromonitoring and frequent ABG collections, we have previously reported that large delta PaCO_2_ in VA-ECMO patients was significantly associated with ICH (OR=2.69; 95% CI=1.18-6.13) but not with composite ABI or mortality(26). Diehl et al. found a PaCO_2_ drop >20 mmHg was associated with mortality but not ABI in VA-ECMO(24). The lack of agreement suggests that peri-cannulation delta PaCO_2_ may be a non-specific phenomenon, and PaCO_2_ management strategies should be further explored in a prospective study with granular ABG data and sufficient sample size. Also, it’s important to highlight that our study focused only on ECPR patients unlike other aforementioned studies.

We systematically investigated the risk factors for ABI, ABI subtypes, and mortality. In addition to pre-ECMO hypoxia and on-ECMO hyperoxia discussed above, lower pre-ECMO pH and requiring renal replacement therapy (RRT) on-ECMO were strongly associated with composite ABI. Pre-ECMO pH has been reported as an independent predictor of survival(27), with pH ≥ 7.0 being associated with better neurological outcome in ECPR patients(28). Especially in ECPR patients with severe metabolic acidosis post-cardiac arrest, pre-ECMO pH is likely a function of the duration of cerebral anoxia/hypoxia prior to the arrest(28). RRT reflects acute renal failure from hemodynamic instability and lack of adequate perfusion, which are also risk factors for ABI(29). In our study, both lower pre-ECMO pH and post-ECMO RRT were also associated mortality. Obesity was also associated with ischemic stroke. Though the pathology underlying obesity contributes to the etiology of ischemic stroke by promoting pro-thrombotic and pro-inflammatory state(30,31), obese patients may also experience delays during cannulation with longer low flow time, which increases likelihood of ischemic ABI(4,32). The main risk factors for ICH were atrial fibrillation and on-ECMO hyperoxia, suggesting that anticoagulant use during VA-ECMO as well as reperfusion injury during ECMO may lead to hemorrhagic conversion of ischemic stroke(33).

The study has several limitations. This was a retrospective study. We did not have access to many Utstein style variables, which may be importantly related to our outcomes of interest. We lacked granular ABG data since the ELSO registry only collects one pre-ECMO and one on-ECMO value, which prevented us from assessing variability or temporal trends (such as rates of change or duration of hyperoxia). The ELSO registry does not collect anticoagulation data, which is an important risk factor of ICH that could not be adjusted in our analysis. Further, as suggested by previous studies, the lack of adjudication of neurological diagnosis and standardized neuromonitoring protocol across contributing centers likely underestimates ABI frequency(34). As the study was retrospective, the association between hyperoxia and ABI is not causal in the absence of information regarding the timing of ABI. Nonetheless, our study contributes significantly to the literature as it utilized a large international cohort that was sufficiently powered to capture clinical practice variations across ECMO centers.

## Conclusions

In a large, international cohort of ECPR patients, we found severe hyperoxia (≥300 mmHg) following ECMO initiation to be associated with acute brain injury and mortality. However, carbon dioxide levels on ECMO or its drop pre-vs. post-cannulation were not associated with composite ABI. Given the risk of reperfusion injury following cardiac arrest, early oxygen delivery practices in ECPR and optimal threshold warrants further study in a prospective trial.

## Data Availability

All data produced in the present work are contained in the manuscript.

## Acknowledgements

None.

