## Supplemental Material for "Arterial Oxygen and Carbon Dioxide Tension and Acute Brain Injury in Extracorporeal Cardiopulmonary Resuscitation Patients: Analysis of the Extracorporeal Life Support Organization Registry"

### Supplemental Figures

**Figure S1.** Unadjusted frequencies of acute brain injury (ABI) by (A) on-ECMO $PaO_2$ and (B) delta $PaCO_2$ category, ischemic stroke by (C) on-ECMO $PaO_2$ and (D) delta $PaCO_2$ category, and intracranial hemorrhage (ICH) by (E) on-ECMO $PaO_2$ and (F) delta $PaCO_2$ category. Negative delta values represent drops in $PaCO_2$ between pre- and on-ECMO initiation, while positive values (">0") represent rises; units are mmHg. Abbreviations – ECMO: extracorporeal membrane oxygenation.

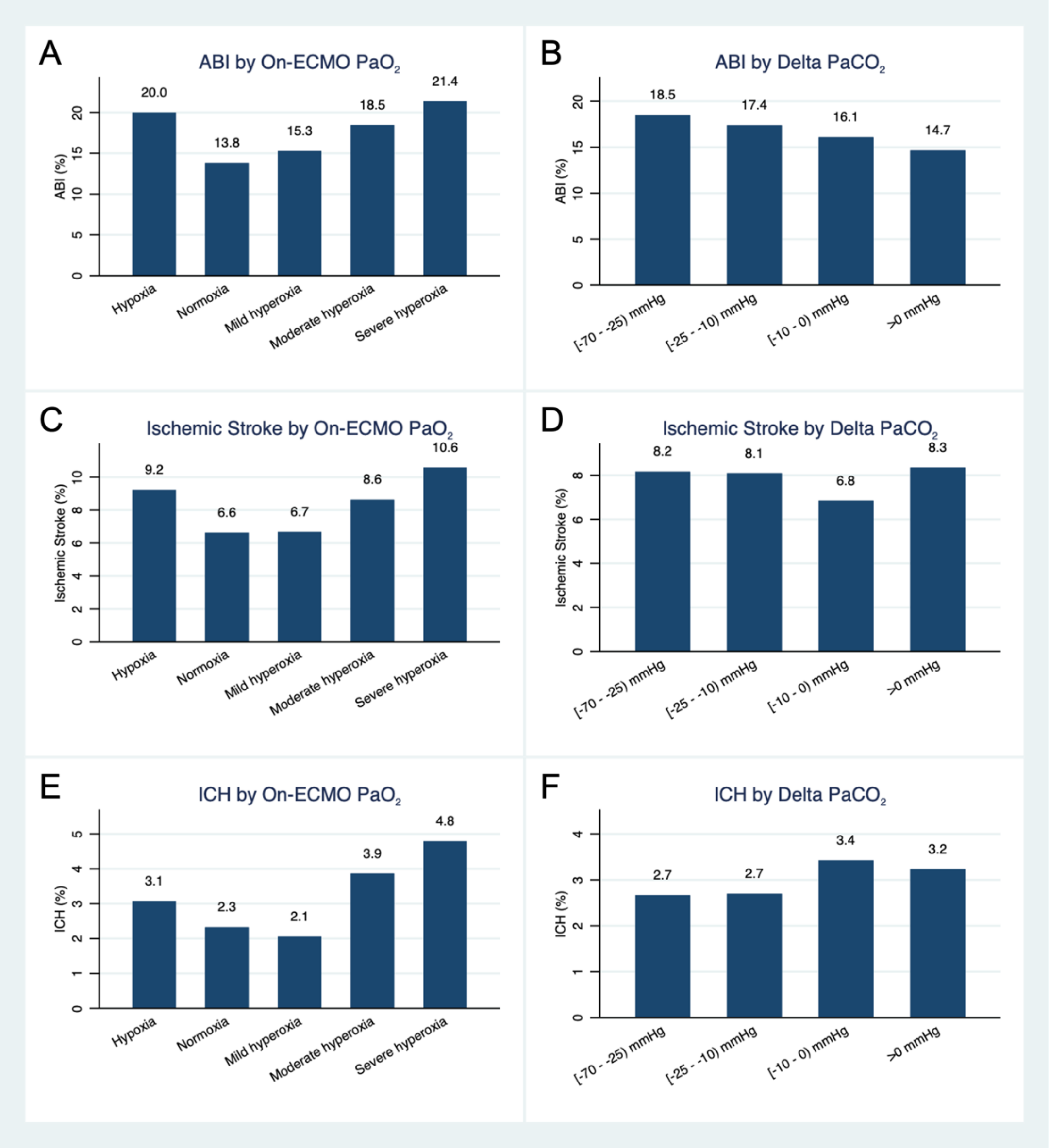

**Figure S2.** $PaO_2$ and $PaCO_2$ parameters by ABI type. ICH: ischemic stroke.

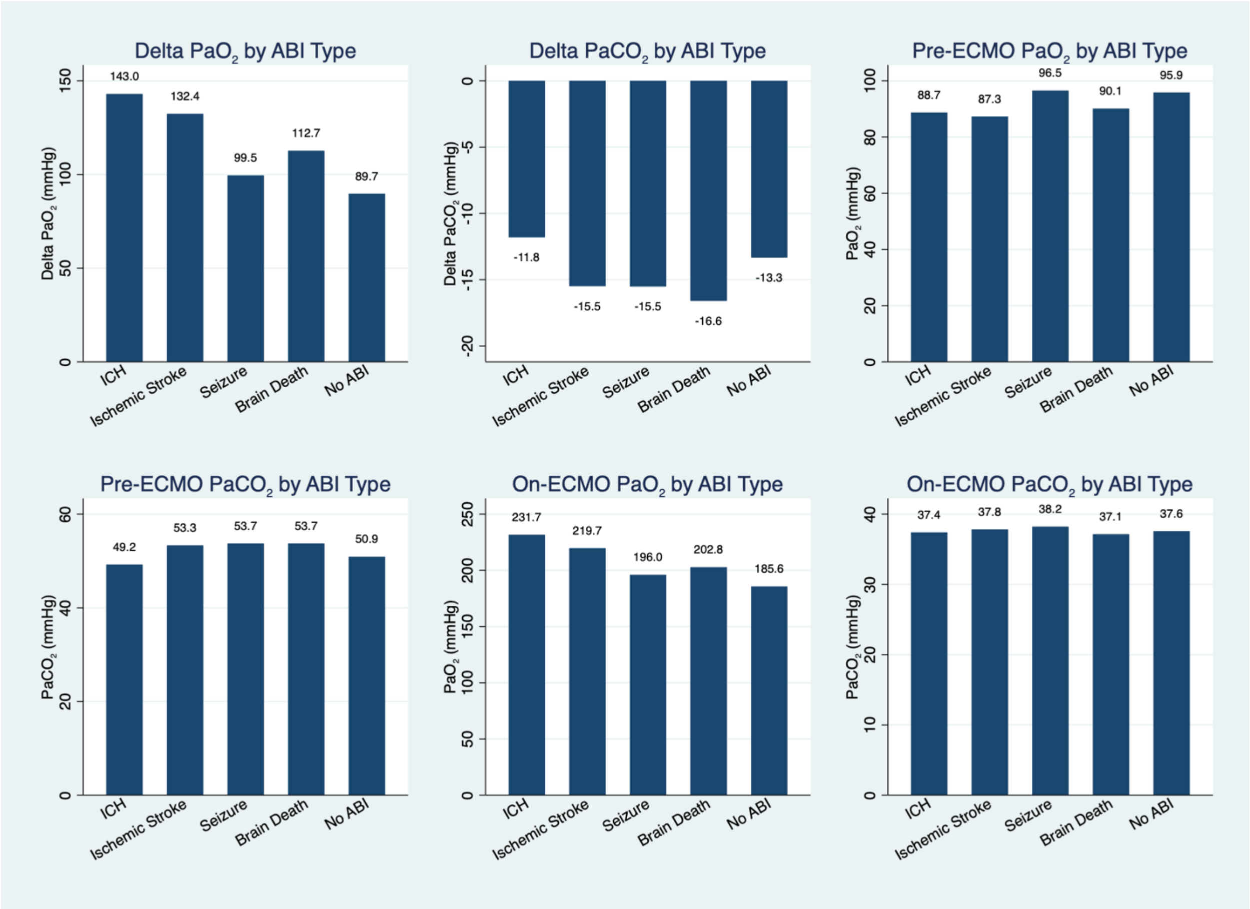

**Figure S3.** Frequencies of mortality by on-ECMO $PaO_2$ and delta $PaCO_2$ category.

ECMO: extracorporeal membrane oxygenation.

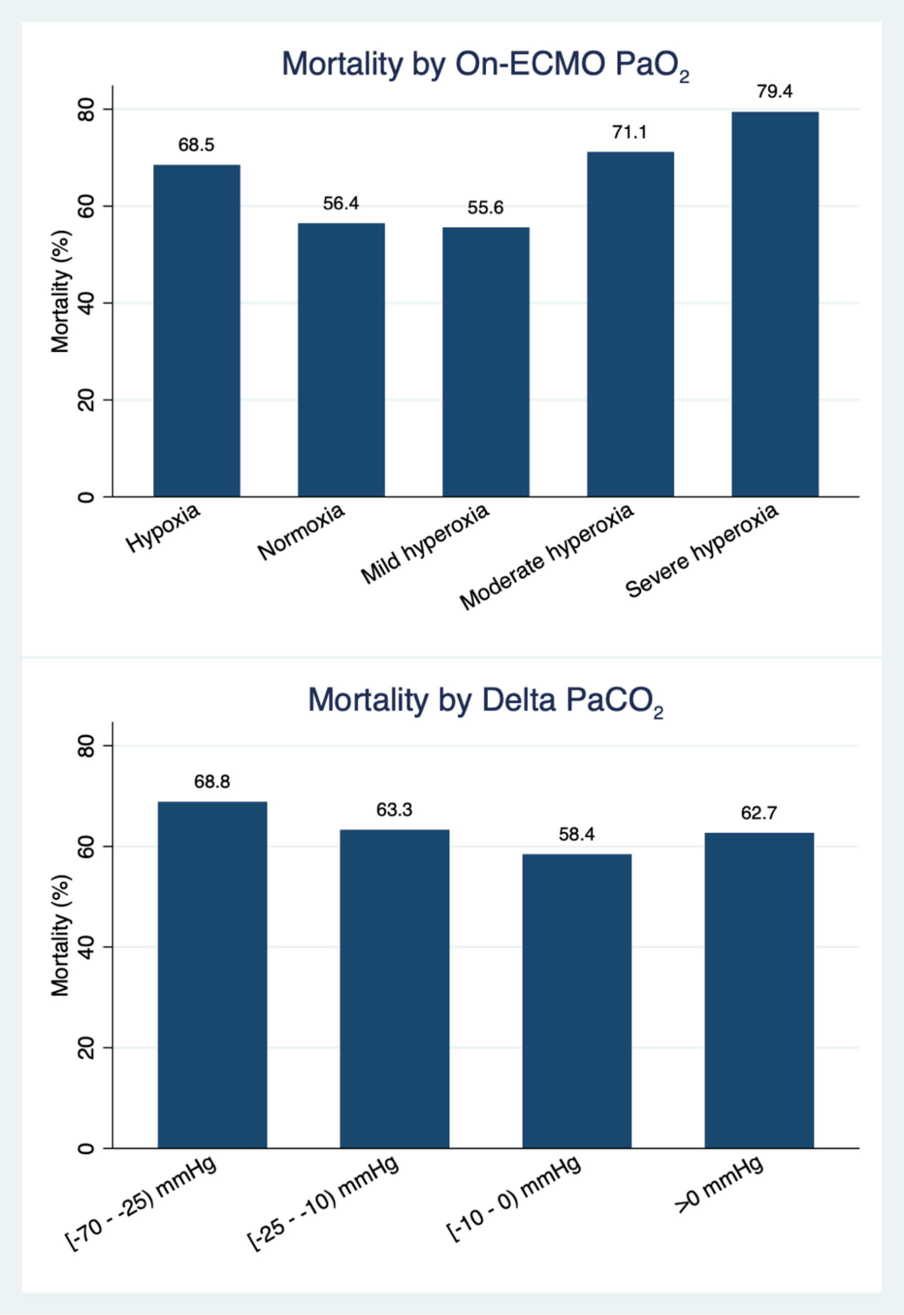

**Figure S4.** Forest plot for multivariable model of mortality. ABI: acute brain injury; MAP: mean arterial pressure; GI: gastrointestinal.

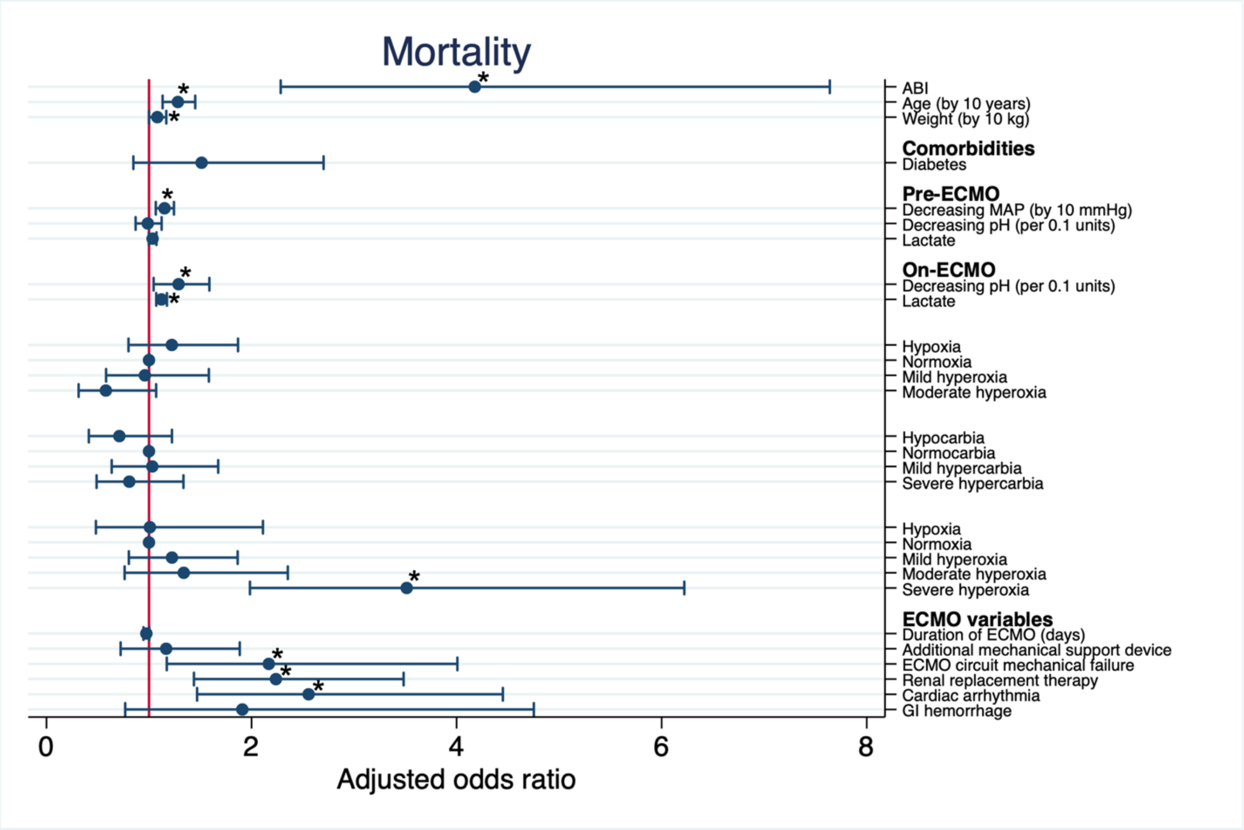

### Supplemental Tables

**Table S1.** Multivariable model for ischemic stroke. C-statistic = 0.70. Variables with a $p<0.05$ are bolded.

| | Multivariable | | |
|---|---|---|---|
| | aOR | 95% CI | p value |
| **Age (by 10 years)** | **0.90** | **0.80-0.99** | **0.04** |
| **Weight (by 10 kgs)** | **1.10** | **1.02-1.17** | **0.008** |
| Race | | | |
| White | Reference | | |
| Black | 1.48 | 0.93-2.37 | 0.10 |
| Hispanic | 1.29 | 0.61-2.74 | 0.50 |
| Asian | 0.81 | 0.50-1.31 | 0.39 |
| Other | 1.52 | 0.94-2.44 | 0.09 |
| Hypertension | 1.61 | 0.98-2.64 | 0.06 |
| **Decreasing pH (per 0.1 units), pre-ECMO** | **1.16** | **1.06-1.28** | **0.001** |
| $PaCO_2$, pre-ECMO | | | |
| Hypocarbia (<35 mmHg) | 1.04 | 0.63-1.71 | 0.87 |
| Normocarbia (35-44 mmHg) | Reference | | |
| **Mild hypercarbia (45-54 mmHg)** | **0.56** | **0.34-0.92** | **0.02** |
| Severe hypercarbia (≥55 mmHg) | 0.77 | 0.51-1.18 | 0.23 |
| $PaO_2$, on-ECMO | | | |
| Hypoxia (<60 mmHg) | 1.49 | 0.76-2.91 | 0.24 |
| Normoxia (60-120 mmHg) | Reference | | |
| Mild hyperoxia (120-199 mmHg) | 0.89 | 0.57-1.37 | 0.59 |
| Moderate hyperoxia (200-299 mmHg) | 1.27 | 0.78-2.05 | 0.34 |
| Severe hyperoxia (≥300 mmHg) | 1.36 | 0.90-2.06 | 0.14 |
| Days on ECMO | 1.02 | 1.00-1.04 | 0.08 |
| **Renal replacement therapy** | **2.03** | **1.47-2.78** | **<0.001** |
| Arrhythmia | 1.27 | 0.86-1.88 | 0.24 |

Abbreviations – ECMO: extracorporeal membrane oxygenation. $PaO_2$: arterial oxygen level; $PaCO_2$: arterial carbon dioxide level. Addition of delta $PaO_2$ and $CO_2$ parameters (both raw values and relative percentages) degraded model performance compared to using only the categorized variables and were excluded from the final model.

**Table S2.** Multivariable model for intracranial hemorrhage. C-statistic = 0.73. Variables with a $p<0.05$ are bolded.

| | Multivariable | | |
|---|---|---|---|
| | aOR | 95% CI | p value |
| **Female sex** | **2.27** | **1.41-3.66** | **0.001** |
| Diabetes | 1.35 | 0.70-2.62 | 0.38 |
| Hypertension | 1.73 | 0.85-3.54 | 0.13 |
| **Atrial fibrillation** | **2.45** | **1.12-5.36** | **0.03** |
| Cardiomyopathy | 1.45 | 0.74-2.85 | 0.28 |
| $PaO_2$, on-ECMO | | | |
| Hypoxia (<60 mmHg) | 1.30 | 0.43-3.94 | 0.64 |
| Normoxia (60-120 mmHg) | Reference | | |
| Mild hyperoxia (120-199 mmHg) | 0.83 | 0.40-1.71 | 0.61 |
| Moderate hyperoxia (200-299 mmHg) | 1.40 | 0.67-2.91 | 0.37 |
| **Severe hyperoxia (≥300 mmHg)** | **1.88** | **1.02-3.47** | **0.04** |
| $PaCO_2$, on-ECMO | | | |
| Hypocarbia (<35 mmHg) | 0.70 | 0.41-1.21 | 0.21 |
| Normocarbia (35-44 mmHg) | Reference | | |
| **Mild hypercarbia (45-54 mmHg)** | **0.39** | **0.15-1.00** | **0.05** |
| Severe hypercarbia (≥55 mmHg) | 1.50 | 0.44-5.15 | 0.52 |
| Days on ECMO | 1.02 | 0.99-1.05 | 0.20 |
| ECMO circuit mechanical failure | 1.42 | 0.79-2.56 | 0.24 |
| Renal replacement therapy | 1.18 | 0.71-1.96 | 0.52 |
| Arrhythmia | 1.62 | 0.91-2.87 | 0.10 |

Abbreviations – ECMO: extracorporeal membrane oxygenation. $PaO_2$: arterial oxygen level; $PaCO_2$: arterial carbon dioxide level. Addition of delta $PaO_2$ and $CO_2$ parameters (both raw values and relative percentages) degraded model performance compared to using only the categorized variables and were excluded from the final model.

**Table S3.** Factors associated with mortality by multivariable logistic regression. C-statistic for multivariable model is 0.81. Variables with a $p<0.05$ are bolded.

| | Multivariable | | |
|---|---|---|---|
| | aOR | 95% CI | p value |
| **ABI** | **4.18** | **2.28-7.64** | **<0.001** |
| **Age (by 10 years)** | **1.28** | **1.13-1.45** | **<0.001** |
| **Weight (by 10 kg)** | **1.08** | **1.00-1.17** | **0.05** |
| Diabetes | 1.51 | 0.85-2.70 | 0.16 |
| **Lower pre-ECMO mean arterial pressure (by 10 mmHg)** | **1.15** | **1.07-1.24** | **<0.001** |
| Lower pH (per 0.1 units), pre-ECMO | 0.99 | 0.87-1.12 | 0.86 |
| Lactate, pre-ECMO (mmol/L) | 1.04 | 1.00-1.07 | 0.05 |
| **Lower pH (per 0.1 units), on-ECMO** | **1.29** | **1.05-1.59** | **0.02** |
| **Lactate, on-ECMO (mmol/L)** | **1.12** | **1.07-1.18** | **<0.001** |
| $PaO_2$, pre-ECMO | | | |
| Hypoxia (<60 mmHg) | 1.22 | 0.80-1.87 | 0.35 |
| Normoxia (60-120 mmHg) | Reference | | |
| Mild hyperoxia (120-199 mmHg) | 0.96 | 0.58-1.58 | 0.87 |
| Moderate hyperoxia (200-299 mmHg) | 0.58 | 0.31-1.07 | 0.08 |
| $PaCO_2$, pre-ECMO | | | |
| Hypocarbia (<35 mmHg) | 0.71 | 0.41-1.22 | 0.22 |
| Normocarbia (35-44 mmHg) | Reference | | |
| Mild hypercarbia (45-54 mmHg) | 1.03 | 0.64-1.68 | 0.90 |
| Severe hypercarbia (≥55 mmHg) | 0.81 | 0.49-1.34 | 0.41 |
| $PaO_2$, on-ECMO | | | |
| Hypoxia (<60 mmHg) | 1.01 | 0.48-2.11 | 0.98 |
| Normoxia (60-120 mmHg) | Reference | | |
| Mild hyperoxia (120-199 mmHg) | 1.22 | 0.80-1.87 | 0.35 |
| Moderate hyperoxia (200-299 mmHg) | 1.34 | 0.76-2.35 | 0.31 |
| **Severe hyperoxia (≥300 mmHg)** | **3.51** | **1.98-6.22** | **<0.001** |
| Days on ECMO | 0.97 | 0.95-1.00 | 0.08 |
| Additional mechanical support device | 1.17 | 0.72-1.89 | 0.53 |
| **ECMO circuit mechanical failure** | **2.17** | **1.17-4.01** | **0.01** |
| **Renal replacement therapy** | **2.24** | **1.44-3.48** | **<0.001** |
| **Cardiac arrhythmia** | **2.56** | **1.47-4.45** | **0.001** |
| Gastrointestinal hemorrhage | 1.91 | 0.77-4.75 | 0.16 |

Abbreviations – ECMO: extracorporeal membrane oxygenation; ABI: acute brain injury; $PaO_2$: arterial oxygen level; $PaCO_2$: arterial carbon dioxide level. Addition of delta $PaO_2$ and $CO_2$ parameters (both raw values and relative percentages) degraded model performance compared to using only the categorized variables and were excluded from the final model.
